# Artificial Intelligence-Enabled Detection of Vascular Perfusion Defects on Ventilation/Perfusion (V/Q) Scintigraphy for Pulmonary Embolism

**DOI:** 10.64898/2026.06.25.26356599

**Authors:** Amir Jabbarpour, Eric Moulton, Sanaz Kaviani, Siraj Ghassel, Wanzhen Zeng, Ramin Akbarian, Anne Couture, Aubert Roy, Richard Liu, Yousif A. Lucinian, Yasser Foufa, Nuha Hejji, Sukainah AlSulaiman, Zeinab Shirazi, Eugene Leung, Ran Klein

## Abstract

Accurate interpretation of planar ventilation-perfusion (V/Q) scintigraphy, used for diagnosing pulmonary embolism (PE) based on PIOPED/EANM guidelines, requires objective assessment of mismatched V/Q defects. Manual delineation of V/Q defects is time-consuming, subject to interobserver variability, and rarely performed in practice, limiting standardized reporting and quantification of disease burden. To address these challenges, we evaluated four modern AI models for automated segmentation of vascular perfusion defects in planar V/Q scans and compared their performance to human annotators. We retrospectively identified 2,118 patients who underwent planar V/Q scans at The Ottawa Hospital (June 2019–February 2023). Six standard projections (ANT, POST, LAO, RAO, LPO, RPO) were included. Four 2D neural networks (U-Net, nnU-Net, Swin UNETR, and a Bottleneck Transformer U-Net [BTU-Net]) were trained on 1,313 patients (7,878 projections) and validated on 329 (1,974 projections) using physician-annotated defects. A hold-out test set of 46 high probability patients was used to evaluate segmentation quality, and defect detection accuracy using free-response receiver operating characteristic (FROC) analysis, where BTU-Net was the only model performing on par with human readers, showing robust sensitivity across the entire range of segmentation probabilities. At 1.5 false positives per projection rate (FPPR), BTU-Net outperformed other models with sensitivity of 0.529 ± 0.026, On a separate hold-out set of low likelihood of disease patients (n=430), the lowest FPPR was 0.08 ± 0.01 for BTU-Net (P<0.0001). BTU-Net enables rapid, consistent, and accurate interpretation of planar V/Q scans. Such tools may enhance diagnostic efficiency, standardize reporting, and support non-expert readers in evaluating PE.

## I. Introduction

Pulmonary embolism (PE) arises most commonly when a thrombus, originating generally in the deep veins of the legs (deep vein thrombosis), migrates through the venous circulation to the heart and ultimately obstructs the small pulmonary arteries(1–6). Ventilation-Perfusion (V/Q) scans constitute one of the main imaging modalities for detecting PE and is primarily favored over other imaging techniques in those patients requiring frequent follow-up imaging and higher sensitivity(7,8). A mismatched V/Q defect—characterized by a hypoperfused region with preserved ventilation—is a hallmark feature of PE(9,10). Although V/Q scans are one of the earliest applications in nuclear medicine, the diagnosis of PE remains both challenging and cognitively demanding. This difficulty arises in part from (1) the existence of multiple interpretation guidelines for V/Q planar(8,11–14) and V/Q SPECT(3,9,15), (2) variability in tracers and imaging protocols—where some centers utilize SPECT, others rely on planar imaging, and some employ both, (3) the requirement for clinicians to visually estimate the extent of perfusion defects as a fraction of bronchopulmonary segmental anatomy, and (4) the high noise and low anatomical information associated with nuclear images. In particular, according to the PIOPED criteria, a high PE probability is indicated by either two large, mismatched defects (>75% of a segment) or two moderate defects (25–75% of a segment) in separate segments.(8) All these factors collectively contribute to substantial interobserver variability.(16)

Artificial intelligence (AI) has been a promising solution for lesion/organ segmentation(17–19), patient level diagnosis(20–22), and prognosis in nuclear medicine and other medical imaging modalities(23–26). Unfortunately, V/Q scans have yet to benefit from modern deep learning architectures such as convolutional neural networks (CNN), vision transformers (VIT), or combinations thereof(27–29) for diagnostic tasks, such as segmenting vascular perfusion defects(5).

This study aims to revitalize AI in the V/Q scan by automatically delineating vascular V/Q mismatches on V/Q planar scintigraphy using modern deep neural networks. In addition, we sought to benchmark each AI model’s performance in relation to typical inter-observer variability of human readers. Inspired by recent work on 3D AI models combining transformers in the bottleneck of a common segmentation architecture, the U-Net, we propose a 2D architecture bottleneck transformer U-Net (BTU-Net) and benchmark it’s performance against established AI models (baseline U-Net, nnU-Net, and Swin UNETR) and nuclear medicine physicians.

## II. Methods

### A. Study Population

All procedures involving human participants were in accordance with the ethical standards of the Ottawa Health Science Network Research Ethics Board (20220303) and with the 1964 Helsinki declaration and its later amendments or comparable ethical standards. This study utilized a retrospective dataset of planar V/Q scans from the VQ4PEDB(30). VQ4PEDB is a large, multi-center—including TOH—database of V/Q nuclear medicine images, primarily created to support AI development for PE diagnosis. It includes over 3,000 annotated V/Q studies along with associated clinical reports, CTPA scans, ultrasound findings, and lab tests, collected from hospitals in Canada and the U.S., with annotations focused on vascular perfusion defects. Patients were categorized according to the modified PIOPED criteria(11), covering a range of diagnostic probabilities for PE including high, intermediate, low, very low, and normal, as well as cases presenting with reverse mismatches between ventilation and perfusion images. For this study, only planar imaging data from the Department of Nuclear Medicine and Molecular Imaging of The Ottawa Hospital (TOH) were included to limit variability in image acquisition methodology.

We retrospectively gathered 1,642 patient studies (9,852 images) having undergone a planar V/Q scan for training and validation. In addition, we devised a holdout test set of 46 patients (276 images) with high probability of PE according to the referring physician’s impression based on modified PIOPED criteria, annotated by six different nuclear medicine fellows and one senior nuclear medicine physician. This hold out set was determined to maximize the number of vascular perfusion defects for downstream model evaluation through pixel level and object-level detection metrics. Finally, a normal holdout test set of 430 patients (2,580 images) with clinical suspicion of PE who were referred to V/Q scintigraphy but ultimately scored as normal modified PIOPED criteria was assembled to quantify the number of false positives introduced by each model on truly normal cases. Overall, this study included 2,118 patients. The study cohort had a mean age of 56 ± 20 years and included 36.1% males and 63.9% females.

### B. Image Acquisition

V/Q scans were acquired at TOH using five different dual-head gamma camera systems: Siemens E.Cam (n=442 patients), Siemens Intevo Evo (n=287), Siemens Symbia (n=445), Siemens Intevo Bold (n=111), and Philips BrightView (n=28). Static imaging was performed starting with the ventilation in the supine position using 370–555 MBq (10–15 mCi) of ^99m^Tc-labeled carbon particles, followed by 148–185 MBq (4–5 mCi) or a reduced dose of 74–93 MBq (2–2.5 mCi) of ^99m^Tc-macroaggregated albumin (MAA) for pregnant or pulmonary hypertension patients for the perfusion. Image acquisition used a ^99m^Tc energy peak of (140 ± 7.5%) keV, across six standard projections on a 256×256 grid with a zoom of 1.46 (pixel size: 1.65×1.65 mm^2^). Anterior (ANT), posterior (POST), left/right anterior oblique (LAO/RAO), and left/right posterior oblique (LPO/RPO) projections were acquired for both perfusion and ventilation portions with 600,000 counts and 150,000 counts per projection respectively. For more information, the reader is referred to the VQ4PEDB paper(30).

### C. Image Annotation

We recruited a total of eight nuclear medicine fellows and a senior nuclear medicine physician to annotate the images used in this study. Each V/Q planar study for the training and validation sets was reviewed independently by one annotator on a first come first serve basis to record their perceived risk of PE using the modified PIOPED criteria and delineate vascular V/Q mismatches across all relevant projections on the V7 Darwin internet hosted platform (V7 Labs, London, UK). All annotations were visually reviewed by the study lead (AJ and in consultation with staff physicians). Studies with obvious annotation errors—including those with missed defects or false positives—were rejected and returned to the annotator with detailed feedback. On average, annotators identified 1.5 ± 3.7 vascular perfusion defects per study calculated across all annotated patients. The average perfusion defect size was 598 ± 472 mm^2^. A hold-out test comprised of 46 patients was annotated by all six nuclear medicine fellows and one senior nuclear medicine physician (7 annotators in total) to provide consensus labels for the downstream clinical validation.

### D. Model Construction

Four CNN models, U-Net, nnU-Net, Swin UNETR, and BTU-Net—designed to segment vascular perfusion defects on V/Q planar scintigraphy (one V and one corresponding P projection at a time) were chosen for this study for the following reasons. The baseline U-Net has long been considered the model of choice for segmentation tasks in medical imaging(31). We used nnU-Net for segmenting V/Q mismatches due to its robust, self-configuring architecture that automatically adapts to the specifics of a given medical imaging dataset. Its state-of-the-art performance across multiple benchmarks, without the need for extensive manual tuning, makes it an ideal choice for handling the variability and complexity inherent in V/Q nuclear scintigraphy data(32). Swin UNETR, a U-Net-like pure transformer learns global and long-range semantic information interaction using hierarchical Swin Transformer with shifted windows as the encoder to extract context features. The BTU-Net architecture offers complementary advantages by integrating a bottleneck transformer that focuses global attention specifically at the network’s deepest layer. We hypothesized that this targeted global context modeling can enhance the representation of complex spatial relationships without significantly increasing computational cost. Therefore, BTU-Net may improve segmentation accuracy by better balancing local detail preservation with global context understanding, especially in challenging cases of subtle V/Q mismatches where fine details and global patterns both matter. As illustrated in Fig 1, the proposed segmentation model is based on a U-Net architecture enhanced with a transformer block in the bottleneck. The U-Net component is a fully convolutional neural network, consisting of a contracting path and an expansive path. The bottleneck includes a transformer block that applies multi-head self-attention with four heads to capture long-range dependencies among features, followed by a feedforward network with two dense layers.

**Fig 1.**
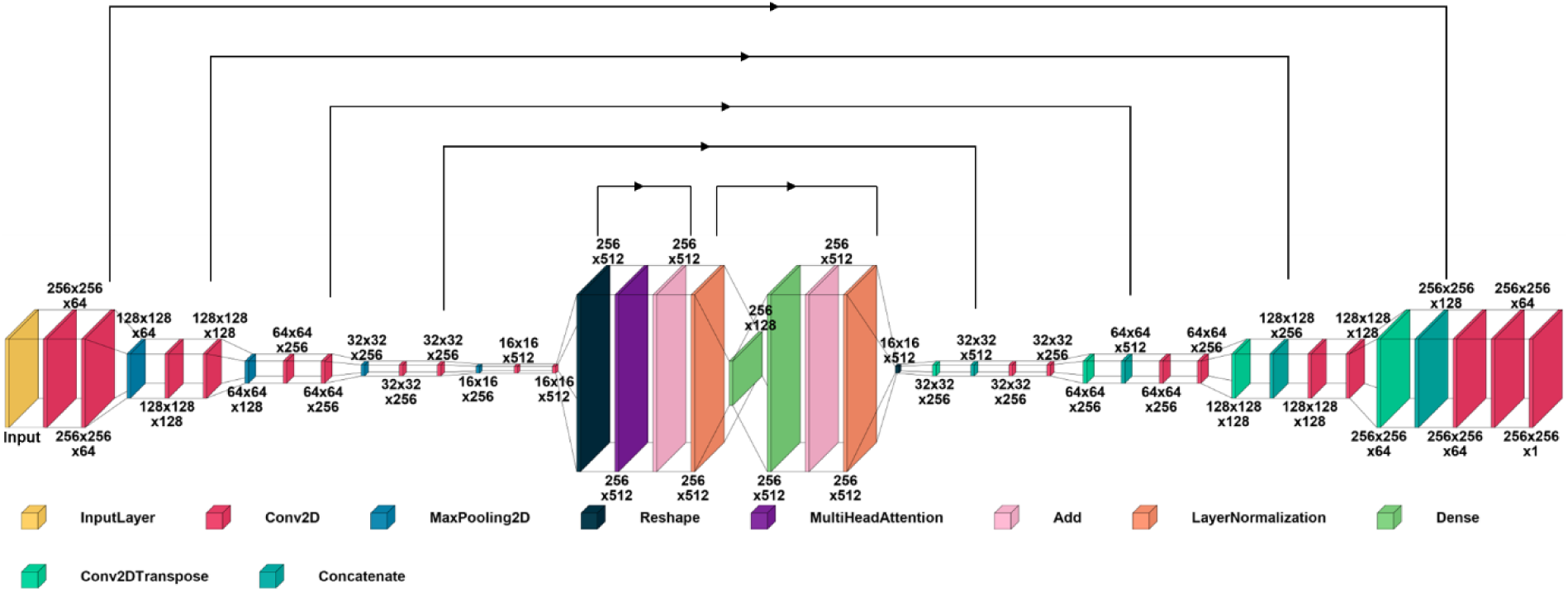
Architecture of proposed 2D BTU-Net Model, a hybrid implementation of Transformers and CNNs for segmenting V/Q mismatched defects.

### E. Model Training

The dataset (n=1,688 studies) was divided into training (n=1,313), validation (n=329), positive holdout (n=46), and negative holdout (n=430). The training and validation sets used a stratified grouping mechanism to prevent data leakage and ensure patient independence of planar projections across splits while maintaining balanced representation across probabilistic PIOPED categories. For the model training set, studies were categorized as follows: 135 high, 151 intermediate, 271 low, 288 very low, 12 Normal Q with V reverse mismatches, and 456 normal cases according to modified PIOPED criteria. The validation dataset categories were 31 high, 46 intermediate, 66 low, 78 very low, 5 Normal Q with V reverse mismatches, and 103 normal cases. In total, the training dataset contained 7,878 two-dimensional projections, while the validation dataset included 1,974 projections.

Prior to model input, ventilation and perfusion images were independently normalized to a [0, 1] intensity range and concatenated along the channel axis to form paired inputs for training and evaluation. U-Net and BTU-Net were implemented in TensorFlow 2.10.1 using Python 3.9.20. Swin UNETR and nnU-Net were implemented using PyTorch 2.3.1, MONAI 1.3.2, and Python 3.11.0. All models utilized He Normal initialization for weights, ReLU activation throughout the networks (except of a sigmoid in the final layer), and an equally weighted combined Dice similarity coefficient (DSC) and binary cross entropy as a loss function. Hyperparameter tuning was performed for all models. All four models were trained using a batch size of 6 and the Adam optimizer with a learning rate of 10^-3^ and a weight decay of 10^-4^. All four models were trained using an early stopping strategy with a patience of 50 epochs and the best-performing model was selected based on the minimal loss function on the validation set. Training was allowed to run for a maximum of 400 epochs.

### F. Evaluation Strategy Across Different Scenarios

Objective evaluation of the predicted mismatched V/Q defects segmentation maps using a single quantitative metric or method is inherently challenging, as the task encompasses multiple interacting factors and complex characteristics. Each projection of V/Q planar scintigraphy could have independent and multiple number of defects. Each projection in V/Q planar scintigraphy can present with no defects, a single defect, or multiple, independent defects, each of which can be correctly identified (true positive), falsely detected (false positive), or missed entirely (false negative). Moreover, in segmentation tasks, calculating true negatives is not informative, as the vast majority of pixels correspond to background (zero probability), rendering this measure unrepresentative of model performance.

For cases with true positive detections, we focused on segmentation overlap and distance-based metrics (elaborated in section II. G.), evaluating segmentation quality only for true positive cases allows for a meaningful assessment of segmentation overlap where defects are actually present. Ground-truth segmentation was defined through the ensemble annotation of seven human readers, by superimposing the delineated mask for each image. The resulting pixel-wise score ranged from zero (excluded by all annotators) to 7 (segmented by all annotators) as demonstrated in Fig 2. A threshold value set at ≥4 annotators (>50%) was the criterion for ground truth segmentation.

**Fig 2.**
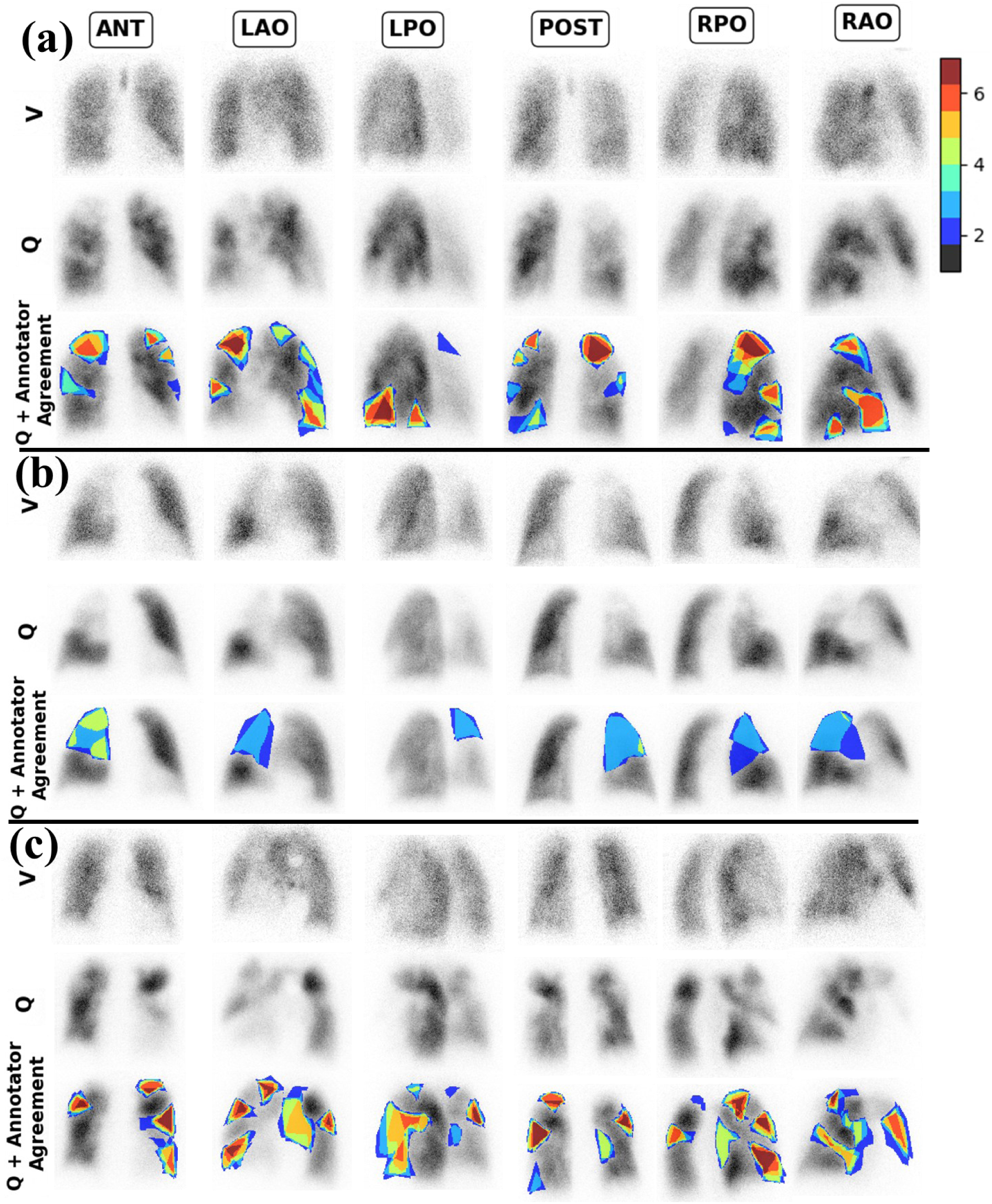
Human annotator variability - Ensemble annotations from seven annotators presented as heat maps for three different patients from the hold-out test set, visualizing magnitude of agreement among physicians at the pixel level.

Since these metrics are not defined in the absence of ground-truth defect segmentations, for projections without annotated defects, model performance was instead assessed using the false positive per projection rate (FPPR), which quantifies the frequency of spurious defect detections in normal images.

Missed vascular defects (false negatives) are inherently captured within the sensitivity dimension of the Free-response Receiver Operating Characteristic (FROC) curve, which characterizes the model’s ability to correctly localize true defects across all projections. Likewise, true positive detections are jointly reflected through FROC sensitivity and Dice, providing complementary perspectives on both detection capability and spatial agreement between predicted and reference defect regions.

### G. Model Segmentation Quality for True Positives

Once a model had been trained, morphological operations were performed to exclude small clusters (< 50 pixels; approximately 3.82 cm²) regarded as noise and not physiologically significant.

We evaluated the quality of generated segmentation masks for true positives by binarizing pixel-wise prediction probabilities at a conventional threshold of 0.5 to create V/Q mismatch defect masks (segmentation). Using connected component analyses, true positives were detected if their centroids lay within a Euclidian distance of 1.65 cm (10 pixels) from the ground truth segmentations. The predicted segmentations for true positives was quantitatively evaluated utilizing the DSC, Jaccard Index, Hausdorff distance (HD), 95^th^ percentile (HD95), and mean surface distance (MSD) against the ground-truth segmentation (ensemble human annotation).

The DSC evaluates the overlap between true and predicted volumes

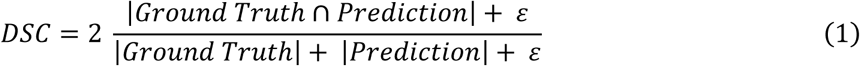

where *ε* = 1 × 10^−6^ was applied to mitigate >1 DSC values in cases with small segments.

The Jaccard Index (a.k.a., Intersection over Union) is another commonly reported similarity metric that quantifies the overlap between two sets:

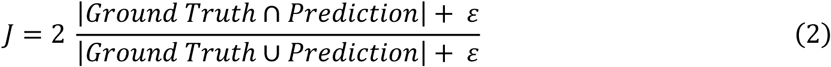

Unlike these overlap-based metrics, the Hausdorff distance (HD) highlights errors at the boundaries. A low Hausdorff distance indicates that the predicted segmentation closely follows the true anatomical boundary. HD in the context of image segmentation, measures how far the predicted boundary (Pred) deviates from the ground truth (GT). Specifically, it captures the largest distance from a point in Pred to the closest point in GT, and vice versa. HD is expressed as follows:

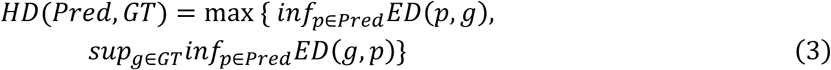

where ED denotes Euclidean distance between two points. Thus, HD reflects the worst-case boundary mismatch between the two sets. Because the HD is sensitive outliers, a modified variant called the 95^th^ percentile HD (HD95) is commonly used. HD95 computes the 95^th^ percentile of the point-wise minimum distances rather than the absolute maximum, making it more robust to noise or minor errors.

Similarly, MSD measures the mean of the shortest distances from each point on the surface of the ground truth to the nearest point on the surface of the prediction, and vice versa, making it a symmetric metric and is expressed as follows:

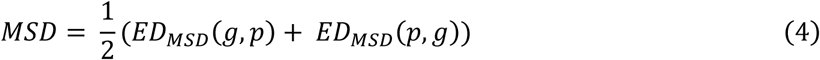

While HD95 represents the region with the worst agreement between segmentation boundaries, MSD represents an average agreement of the entire boundary.

Since the number and spatial distribution of predicted connected components (i.e., true positives) vary across models, direct comparison of segmentation quality and formal statistical testing are not straightforward. Therefore, no statistical analysis was performed for these metrics.

### H. Clinical Evaluation: Model Free Response Characterization

As mentioned, to benchmark the model’s ability to detect vascular perfusion defects against human readers, a subset of included annotators (N=7) was tasked to segment vascular perfusion defects on the 46 high probability PE patients of the hold-out test set, and the ground truth was defined as pixels segmented by ≥4 as V/Q mismatch defects.

FROC curves were generated considering pixel-wise segmentation probabilities as operating points. Then, using connected component analyses, candidate segmentations of vascular defects were considered true positives if their centroids lay within a Euclidian distance of 1.65 cm (10 pixels) from the centroid of the ground truth segmentations. This process was performed to generate a FROC curve for each model against each physician, and the 95% confidence intervals (CI) were derived for each model by bootstrapping all FROC curves across the seven annotators. Mean sensitivities and CIs of individual annotators were calculated against all six other annotators. These served as clinical reference against which AI model performance could be evaluated. The resulting FROC curves depict the trade-off between sensitivity for the detection of true mismatch regions and the rate at which false positives (reporting mismatch regions that are not real) are produced. This trade-off is dependent on the threshold used to classify whether the pixel-wise probabilities produced by the respective model correspond to a defect region versus background. Using these curves, we also reported model sensitivities at common preset FPPR’s of 0.5, 1.0, and 1.5. These in turn can be used to select a task-specific operating threshold for the model in the context of a clinical application (e.g. screening versus decision to treat).

### I. False positives on Normal Studies

The FPPR on normal studies assigned as normal PIOPED score was evaluated on studies with no visible defects as reported both clinically and by annotators (n = 430 studies; n = 2580 projections). A false positive was defined as the presence of any predicted connected component with >50 pixels exceeding a probability threshold of 0.5 in these negative cases. Pairwise comparisons between models were subsequently performed using two-sample proportion z-tests to identify statistically significant differences in FPPR.

## III. Results

### A. Variability in Image Annotations

Overall, we observed considerable variability among human annotators, particularly in the identification of peripheral and small subsegmental defects and in cases where both ventilation and perfusion were heterogeneous. Disagreement was also common in regions with decreased, but not absent, perfusion signal, as well as in cases with partial mismatches between ventilation and perfusion. These findings highlight differences in defect interpretation across annotators. Fig 2 demonstrates heatmaps of the number of annotators that segmented a pixel as belonging to a V/Q mismatch defect in three representative patients. With red regions indicating consensus of presence of defect and no color (gray scale) indicating consensus of absence of defect, and blue to orange regions corresponding to varying amounts of inter-annotator variability. Case (a) indicates relatively good agreement in segmentation amongst the 7 annotators. Case (b) illustrates poor agreement in both the identification of defects and the assessment of their extent. Case (c) shows moderate discrepancies in defect localization.

Fig 3 summarizes the five segmentation performance metrics for true positive segmentations by the four models, with respect to the ensemble of all human annotators as the ground truth. These results consistently demonstrated better mean performance ranking (higher DSC and Jaccard, and lower MSD, HD95 and HD) for BTU-Net followed by Swin UNETR, nnU-Net, and U-Net. BTU-Net consistently tended towards higher overlap metrics and lower distance-based errors compared to the other models. BTU-Net median (min-max) scores were DSC= 0.72 (0.00–0.93), Jaccard=0.57 (0.00–0.88), MSD=2.77 (0.53–18.03) mm, HD95=14.74 (3.30–66.51) mm and HD=8.87 (1.65–55.97) mm.

**Fig 3.**
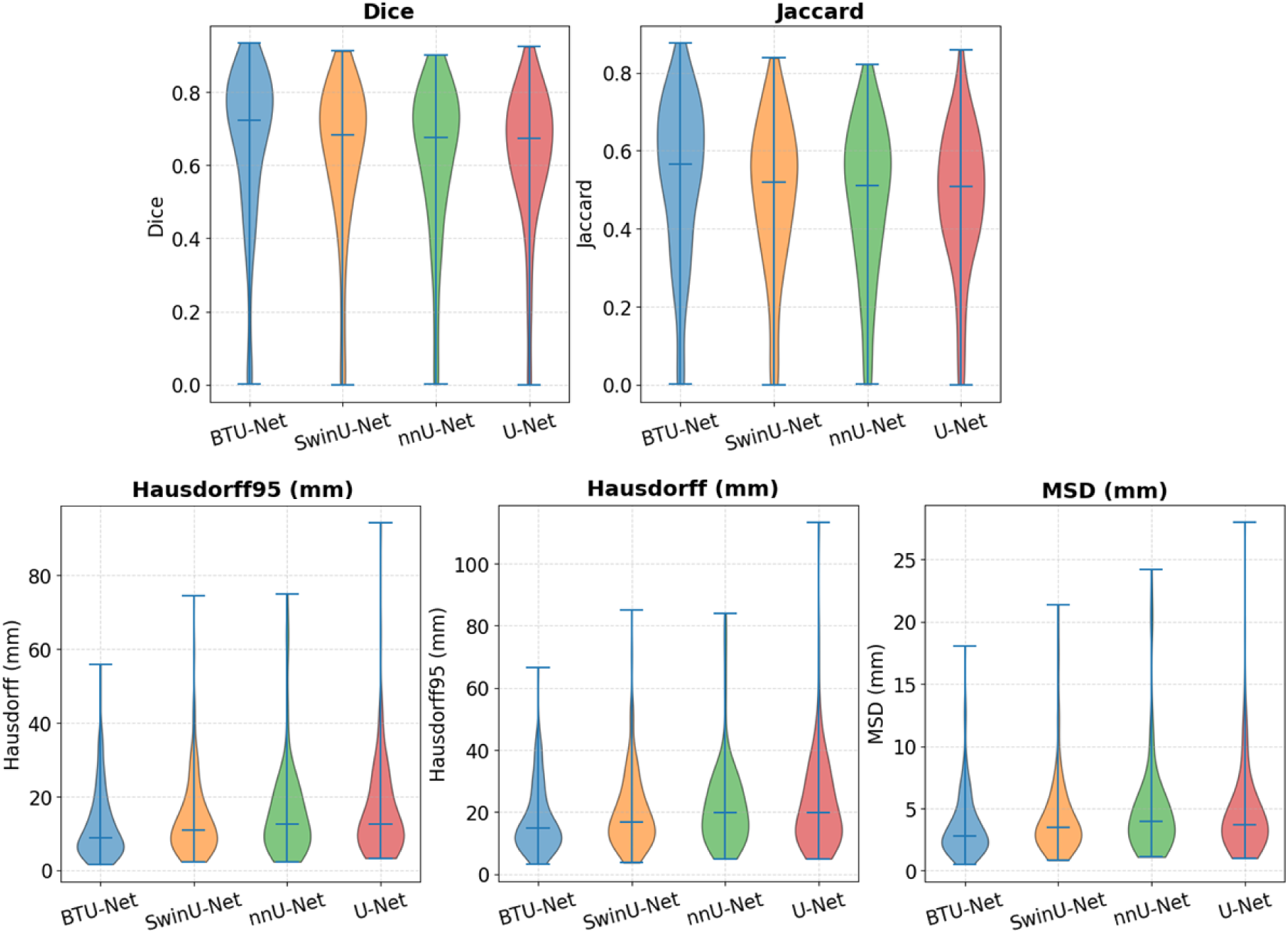
Violin plots of segmentation performance metrics for each model, including dice score, Jaccard index, Mean Surface Distance (MSD), 95^th^ percentile Hausdorff Distance, and Hausdorff Distance. Only true positive predicted defects are included.

### B. Clinical Evaluation: Model Free Response Characterization

Superimposed on Fig 4 are the FROC analyses of the four models, which also shows the sensitivity versus FPPR of each annotator in comparison to each other. FPPR here presents number of predicted false positives per projection rate on abnormal patients (n=46 studies, 276 projection images). The FROC curves revealed that BTU-Net is the only model performing on par with human readers as indicated by overlapping confidence intervals with the human annotators’ sensitivities and false-positive rates.

**Fig 4.**
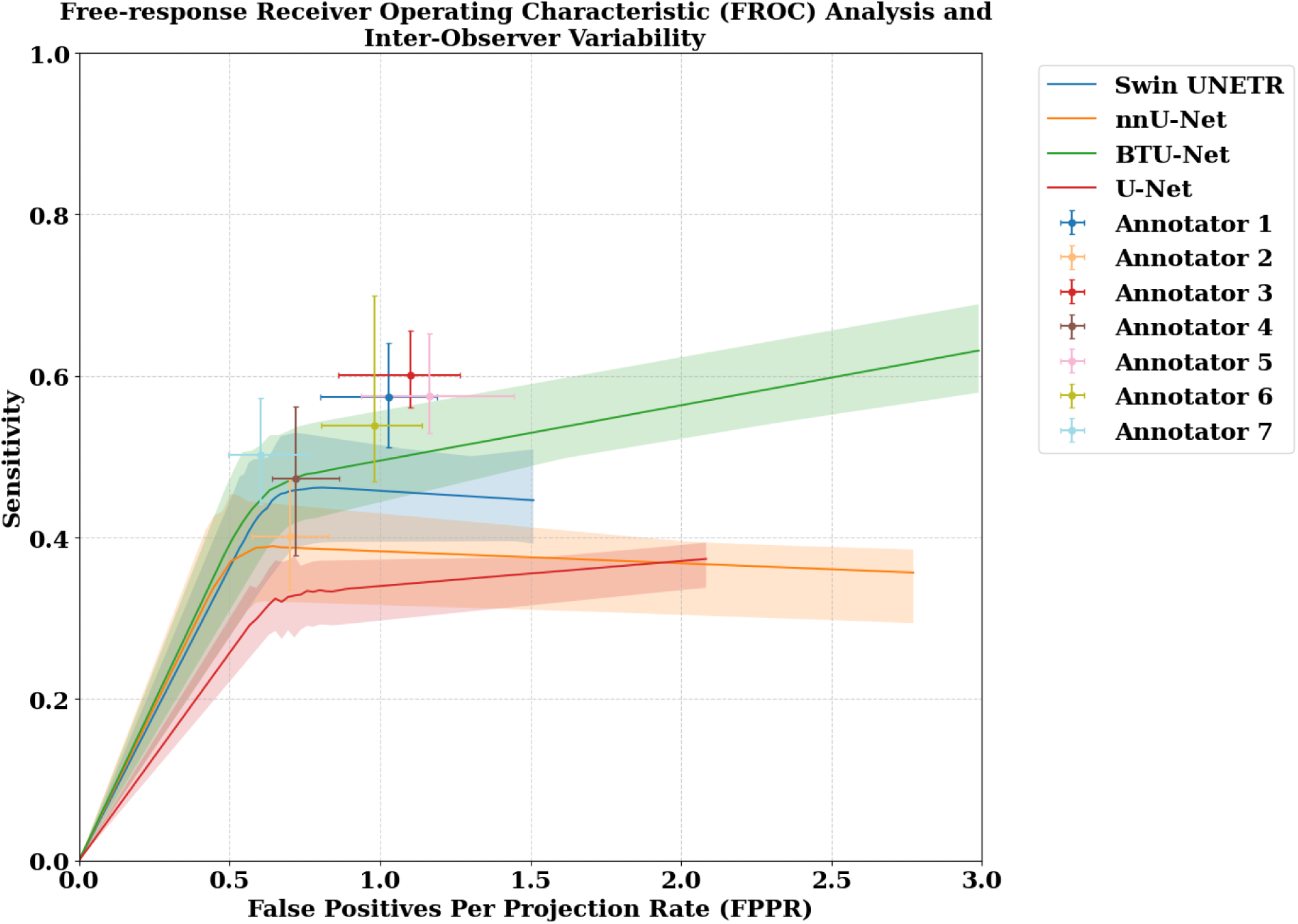
FROC curves with respective confidence intervals for U-Net, nnU-Net, Swin UNETR, and BTU-Net evaluated on holdout test set. Mean sensitivity and confidence intervals of individual annotator were calculated against six other annotators. Confidence intervals of each model were measured in respect to all annotators.

Nevertheless, it does not achieve high sensitivity (<60%) even at 2.5 FPPR. Likewise, none of the human readers exceeded 60% sensitivity on the same set of images. Table 1 summarizes model performances at key operating points of 0.5, 1.0, and 1.5 FPPR. Corresponding to Fig 4, BTU-Net achieved the highest sensitivities with 0.391 ± 0.042, 0.495 ± 0.029, and 0.529 ± 0.026, while the second best performing model was Swin UNETR with 0.363 ± 0.043, 0.458 ± 0.031, 0.446 ± 0.030 at the same FPPRs.

**Table I.** Lesion detection mean sensitivity ± standard deviation at 0.5, 1.0, and 1.5 FPPR.

| Network | Sensitivity @ FPPR = 0.5 | Sensitivity @ FPPR = 1.0 | Sensitivity @ FPPR = 1.5 |
| --- | --- | --- | --- |
| BTU-Net | 0.391±0.042 | 0.495±0.029 | 0.529±0.026 |
| Swin UNETR | 0.363±0.043 | 0.458±0.031 | 0.446±0.030 |
| nnU-Net | 0.368±0.044 | 0.383±0.029 | 0.376±0.026 |
| U-Net | 0.257±0.020 | 0.340±0.019 | 0.355±0.015 |

It is worth noting in Fig 4 that human annotators operated at approximal FPPR between 0.6 and 1.2; hence FPPR=0.5 and 1 represents convenient operating points for comparing AI and human annotator sensitivities. Sensitivity also varied substantially across annotators, reflecting considerable inter-observer variability in interpretation. Across annotators, sensitivities ranged from 0.401 to 0.502 at FPPR ≈ 0.5, and from 0.539 to 0.601 at FPPR ≈ 1.0. Nevertheless, no annotator revealed a clear advantage (closer to the top left corner) over other annotators. Rather, these results demonstrated that certain annotators favor sensitivity to V/Q mismatch defects at the expense of more false positives or vice versa.

Based on the FROC analysis it was determined that model performance did not vary significantly at intermediate values of the operating point (threshold pixel class probability by the models).

### C. False positives on Normal Studies

On normal studies (n = 430 studies; n = 2,580 projections), the mean ± standard deviation number of false positive per image for each model was as follows: BTU-Net, 0.08 ± 0.01; Swin-UNETR, 0.12 ± 0.01; nnU-Net, 0.22 ± 0.01; and U-Net, 0.15 ± 0.01, demonstrating the superiority of BTU-Net over all other models (P<0.0001).

### D. Qualitative Assessment of Model Performance

To complement the quantitative evaluation metrics, qualitative assessments were conducted using representative examples from the hold-out test dataset. Fig 5 illustrates segmentation outputs from each model on a challenging case selected from hold-out test set to reflect a range of defect complexity and anatomical variability. This case proved similarly challenging for human readers (Fig 2.c). Likewise, in Fig 6 a patient without defects is shown. For each patient, model predictions are displayed alongside the corresponding ground truth annotations to enable visual comparison. The baseline U-Net frequently predicted anomalously large and irregularly shaped segments in areas of vascular perfusion defects, often merging multiple distinct defects into a single region. In doing so, it incorrectly encompassed normal lung tissue while also missing several clinically relevant defects. These issues were somewhat mitigated in the predictions of nnU-Net and Swin UNETR. While Swin UNETR demonstrated fewer instances of missing clinically important defects compared to U-Net and nnU-Net, it still produced imprecise segmentations that were not consistent with the wedge-shaped defects expected from anatomical lung segments. These shortcomings were generally (but not always) addressed by BTU-Net, which generated more accurate and anatomically plausible predictions, showing best agreement with human annotator concordant regions.

**Fig 5.**
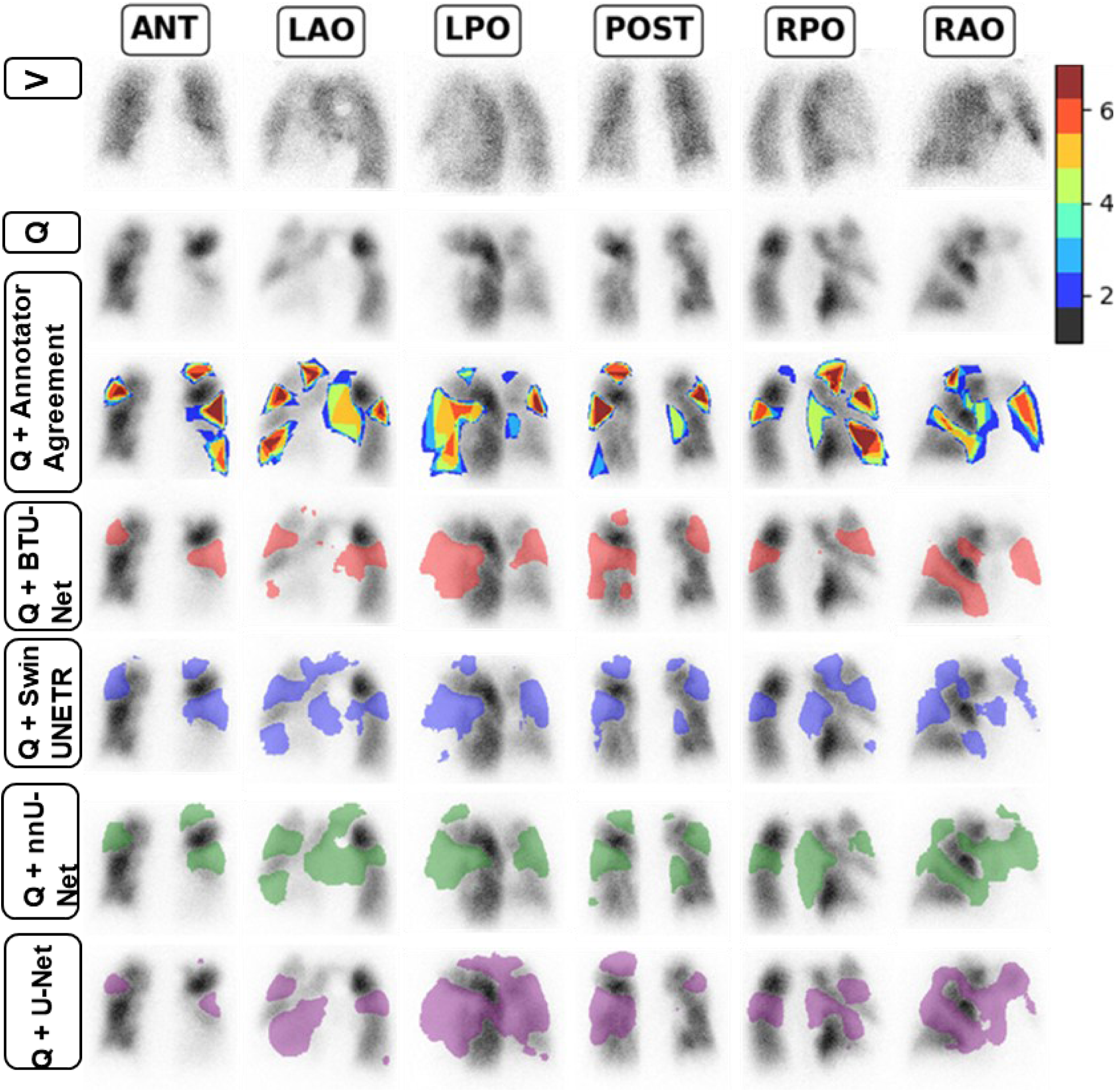
A challenging case illustrating variable segmentation of V/Q mismatch defects both by human annotators and models.

**Fig 6.**
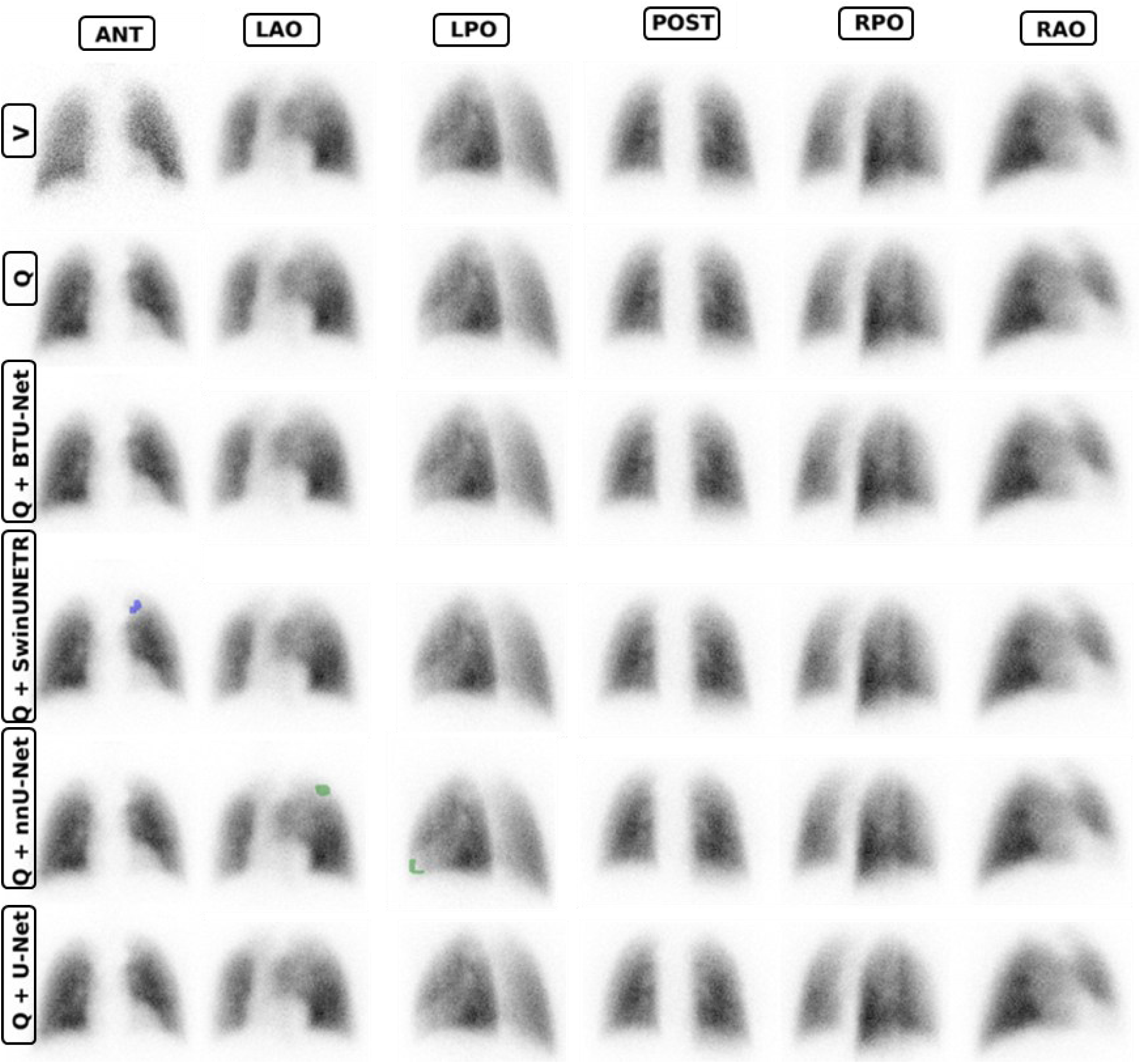
Qualitative comparison of segmentation outputs across models on a normal (defect-free) case. Unlike other architectures, BTU-Net did not introduce false positive defects, demonstrating superior specificity on normal patient data.

## IV. Discussion

In this study, we developed a novel V/Q planar mismatched segmentation approach based on deep learning approaches. The models were trained on ground truth labels provided by 9 nuclear medicine physicians, capturing a broad range of interpretation strategies. Of these models, the BTU-Net architecture was intentionally designed to combine the spatial hierarchies captured by the U-Net with the global attention capabilities of transformers, enhancing segmentation performance by leveraging both local and global contextual information. This design enabled BTU-Net to be the only model that performed on par with human readers (despite being trained on the same data as the other models).

Our claimed study complies with guidelines of AI Task Force of SNMMI including ‘Nuclear Medicine and Artificial Intelligence: Best Practice for Algorithm Development’(33) and ‘Nuclear Medicine and Artificial Intelligence: Best Practices for Evaluation (the RELAINCE Guidelines)’(34) for developing BTU-Net and clinical-task-based evaluation of proposed model. As encouraged by these guidelines, we validated the performance of BTU-Net against three state-of-the-art architectures—Swin UNETR, nnU-Net, and U-Net—representing varying levels of complexity as well as against human readers. The BTU-Net model had better segmentation performance, superior detection performance, and lower false positive rates. Hence it was judged to have the best potential for clinical implementation and as a training tool for novice nuclear medicine practitioners.

This study expends on our previous abstract (35) and is unique owing to novel models and application for lung V/Q mismatch analysis, the diversity and large size of the training data sourced from multiple expert annotations, evaluation of both defect segmentation and detection, and achieved diagnostic accuracy approaching that of trained nuclear medicine physicians for detecting vascular perfusion defects.

### A. Prospects of a PE assessment tool

To the best of our knowledge, this is the first attempt to directly segment vascular perfusion defects on V/Q images (a) by humans and (b) evaluate interobserver variability on defect level using deep learning. Prior studies have primarily focused on either identifying PE or predicting PIOPED score mostly using radiomics and shallow artificial neural networks (ANNs) with feature engineering in suspected PE cases using V/Q scintigraphy(5,36–41). Our work represents only the second study on modern AI applied to PE detection in V/Q imaging since 2008, following a 17-year gap in literature as highlighted by a recent literature review from our team(5). The only other recent study to incorporate a modern attention mechanism focused solely on classifying perfusion defects into four broad categories: grade 0, no or insignificant defect; grade 1, localized peripheral defect; grade 2, defect involving up to one lobe; and grade 3, defect exceeding one lobe. However, that approach did not include segmentation of vascular defects or any clinical evaluation. Instead, it functioned as an opaque tool to assist with PIOPED scoring by categorizing V/Q images based on the extent of subsegmental, segmental, or lobar defects. In future work we aim to similarly classify V/Q studies from our segmented V/Q defects with the aim of achieving a mode explainable model.

### B. Clinical Evaluation in the Context of Interobserver Variability

A large degree of interobserver variability is demonstrated in Fig 2, depicting annotation discrepancies for three high probability patients. This finding explains the large variability in human observer performance (Fig 3) as manifested by sensitivities ranging between roughly 40 and 60% and FPR ranging between 0.6 and 1.2 per projection image (FPPR). Previous literature has reported similar large interobserver variability in reporting V/Q studies, albeit in SPECT (16). Most inconsistencies arise from disagreements in segmenting V/Q mismatched defects (Fig 2a, ANT, LAO and POST; Fig 2b) as well as in determining the overall extent of defects (Fig 2a, b, and c).

Figs 2.c and 5 exemplify a particularly challenging case with heterogenous perfusion distribution leading to inconsistent segmentation between human readers and between models respectively. This qualitative comparison is both challenging and subjective. In contrast, in Fig 4, by superimposing the sensitivities and FPPR of human readers in comparison to one another with the FROC curves of machine observers, we were able to perform an unbiased comparison of machine observers in the context of current clinical performance on the task of PE mismatch detection. The above-described discrepancies in human-derived labels from V/Q image interpretations—considered the second-best gold standard—may in fact be the primary limitation preventing models from achieving higher sensitivity on the FROC curve. Pulmonary angiography, the gold standard for diagnosing PE has virtually vanished due to its invasive nature, as noticed by Gottschalk and colleagues(12) leaving deep learning for PE with a great challenge for identifying real defects. This subsequently gives rise to significant interobserver variability as observed with our FROC analyses of human readers in Fig 4. Labeling data has been also clearly emphasized by Bradshaw et al. in the AI Task Force of the Society of Nuclear Medicine and Molecular Imaging, identifying it as the component with the “greatest dividends”(33). Regardless, the variability of human annotators and their use as a reference truth undermines the ability to both train and evaluate the performance of machine observers.

Another contributing factor to discrepancies in human annotations is the variability in interpretation guidelines across centers for V/Q SPECT and planar imaging, as well as the influence of individual gestalt impressions incorporated into the final diagnosis. Also, some physicians are trained primarily in one modality over the other. Incorporating delineation of segmental and subsegmental defects (aided by AI as developed herein) into the clinical workflow could help train novice clinicians regarding these anatomical structures and facilitate communications of perfusion defect findings. With more common segmentation, better consensus amongst clinicians may emerge to aid training and validation of future AI.

### C. Performance Metrics

Herein, we evaluated the models using three distinct criteria: defect segmentation quality, clinical defect detection, and false positives in normal cases. Segmentation similarity metrics primarily focus on the quality of the predicted segmentation masks—specifically, the overlap between ground truth and predicted regions—only for true positives. The clinical defect detection through FROC analysis evaluates the clinical relevance of the predicted defects. Finally, the FPPR on normal studies measures each model’s propensity to generate artifactual defects in physiological normal projections, providing a stringent assessment of its robustness and clinical trustworthiness. Although FPPR analysis was conducted exclusively on cases reported as normal clinically and by at least one annotator, it is important to note that these patients were still clinically suspicious and had been referred for V/Q imaging.

The inclusion of three evaluation strategies is associated with the clinical tasks performed towards PIOPED criteria scoring which is dependent on both number of defects and their sizes. Furthermore, the segmentation quality is an important element towards explainability of a PIOPED score. A key limitation of the DSC, Jaccard, HD, HD95 and MSD is their inability to reflect the number of missed defects for each V/Q scan by a model. Another important drawback of all these segmentation metrics is the lack of gold standard against which the results can be compared.

We therefore complimented our evaluation for the clinical task of defect detection, by counting true positives (TP) and false positives (FP) and false negatives (FN). The 10-pixel threshold was arbitrary choice for FROC analysis to allow inclusion of smaller predicted defects relative to the bigger corresponding ground truth segmentation. Predicted segmentation maps were variably thresholded based on pixel-wise probability values to construct FROC curves. Given the absence of a standardized framework for FROC analysis across segmentation tasks and imaging modalities, methodological variations are to be expected and often tailored to the specific clinical or research context.

Finally, to assess fidelity on normal patients we evaluated the models on 430 studies deemed normal in clinic and by annotators. While the models did predict some defects, these were small and would likely be dismissed in the clinical context as noise or not clinically significant. Alternatively, these could be perceived as real defects that were not annotated for the same reasons.

### D. Study Level versus Image Level Interpretation

In this work, we evaluated the detection of V/Q mismatch defects on a per image basis. However, the clinical question concerns the interpretation of the state of the patient. Hence the 6 projections offer redundant information that may be used to refute or support a suspected defect on any one image. Developing a methodology to consolidate the 6 projections to a study level interpretation was beyond the scope of this work. Nevertheless, a human reader that would digest machine predicted defects would compare the projections and is therefore likely to reduce the study level false-positive rate associated with defects that are not reproduced between projections, with likely minimal impact on the sensitivity. Hence selecting a high sensitivity operating point (low pixel-wise probability threshold), at the expense of elevated FPPR, may be recommended for this application, but further validation would be required.

### E. Other Applications and Pathologies

Image quality assessment plays a critical role both in clinical workflows and during the development of machine learning models for translating and reconstructing medical images. While widely used full-reference metrics like PSNR and SSIM have proven effective in natural image tasks, their limitations in medical applications have been repeatedly documented, pointing to a disconnect between algorithmic development and real-world clinical relevance. This is particularly significant because conventional loss functions and metrics primarily assess image similarity based on general criteria, often failing to fully capture the preservation of clinically relevant information and pathological defects encoded within the image(42,43). As recommended by Herath et al.(43), image quality should be evaluated based on task-relevant downstream performance. In this context, BTU-Net, as an objective reader, holds promise to be leveraged in training and evaluating various AI models aimed at enhancing the quality of V/Q scans in such a way that accurately preserves the clinical presentation of defects. By acting as an AI-based objective reader, BTU-Net could serve both as a clinical loss function during model training and as a robust evaluation tool of V/Q scintigraphy count enhancement. This would enable more meaningful comparisons between AI-enhanced images, ensuring that crucial clinical information is retained and effectively represented.

When a V/Q planar interpretation yields intermediate probability for PE, the scan is considered non-diagnostic and is referred to CTPA for further investigation, wherein a patient will be exposed to 5-30 times higher dose of radiation(44,45). BTU-Net could serve as a valuable tool to enhance diagnostic confidence for physicians and elevating non-expert to expert reader level, potentially reducing redundant imaging and minimizing unnecessary referrals for CTPA. Nevertheless, due to the relative ease of CT procedures and the widespread availability of CT technology, CTPA will continue to play a crucial role in emergency settings and in situations where nuclear V/Q scintigraphy is not accessible.

The proposed AI model and VQ4PEDB developed for V/Q mismatch segmentation could be readily reconsidered to benefit diagnosis of other pathologies, such as chronic thromboembolic pulmonary hypertension (CTEPH). V/Q scintigraphy remains the gold standard for screening CTEPH, a curable cause of pulmonary hypertension that is often underdiagnosed. Recent studies have established that using a threshold of at least 2.5 segmental mismatched perfusion on planar V/Q scans provides optimal diagnostic accuracy for CTEPH, achieving 100% sensitivity and over 94% specificity(46). VQ4PEDB(30) included a mix of patients who underwent V/Q scintigraphy at both acute and chronic stages, making our model a strong candidate for diagnosing CTEPH in suspected cases with minimal need to modify BTU-Net.

This study establishes a foundational benchmark for the segmentation of V/Q mismatched defects – revitalizing previously neglected area of AI in nuclear V/Q pulmonary imaging – achieving performance on par with expert human annotators. Looking ahead, it will be critical to further refine segmentation accuracy, integrate predictive modeling of PIOPED criteria, and harness the enhanced anatomical separation offered by V/Q SPECT imaging. Equally important is the development of sophisticated registration techniques that can accurately map predicted defects onto a bronchopulmonary atlas(47), thereby enabling the generation of robust, objective metrics for quantifying PE burden. Collectively, these advancements will not only enhance the precision of automated analysis but also solidify its role as a valuable adjunct in clinical decision-making.

### F. Limitations and Future Works

Despite our promising results, our work still presents certain limitations. Subsegmental defects remained difficult to accurately detect. Therefore, our model should still be used as a diagnostic aid with a human in the loop. These readers must still be well versed in interpreting planar V/Q studies so that they can assess the accuracy of the V/Q mismatch defect detection and agreement between complimentary views towards a consolidated interpretation. Nevertheless, these predictions may be useful as a guide as a decision support tool and as a second reader to reduce missed defects.

This single center study did not evaluate the generalizability to other image variants including matrix size, ventilation agents, noise levels (image counts) and pseudo planar projections from SPECT. Additional testing on representative data is essential before clinical application. Likewise, techniques standardizing image characteristics may also aid generalizability. By example resizing of scintigraphic image grids, as highlighted by a recent study from our team through Poisson noise-based resampling(48) may be beneficial to enhancing generalizability.

## V. Conclusion

We developed and compared several AI for automatic segmentation of vascular perfusion defects on V/Q planar scintigraphy, an underutilized and largely overlooked nuclear medicine imaging modality in AI applications. We validated our model against other state-of-the-art models and physicians, demonstrating that bottleneck U-Net neural network, a novel hybrid architecture combining CNNs and transformers, can perform on par with expert readers.

## Data Availability

Reasonable data requests can be directed to the senior author.

## Declarations

### I. Ethics Approval

All procedures involving human participants were in accordance with the ethical standards of the Ottawa Health Science Network Research Ethics Board (#20220303) and with the 1964 Helsinki declaration and its later amendments or comparable ethical standards.

### II. Consent for publication

N/A

### III. Availability of data and material

Reasonable data requests can be directed to the senior author.

### IV. Competing interests

Amir Jabbarpour is co-funded by Jubilant DraxImage Inc. Eric Moulton is an employee of Jubilant DraxImage Inc. Ran Klein received revenue shares and is consultant to Jubilant DraxImage Inc. for Rubidium-82 generators and elution systems. Ran Klein performs collaborative research and receives in-kind support from Hermes Medical Solutions and Invia Medical Solution. Ran Klein consults to Boston Scientific.

## I. Funding

This work was supported by MITACS through the MITACS Accelerate program (grant no. IT29092) and by INOVAIT (project number 2022-4041). Funding for this project was provided in part by INOVAIT through the Government of Canada’s Strategic Innovation Fund. Ce projet a été financé en partie par INOVAIT dans le cadre du Fonds stratégique pour l’innovation du gouvernement du Canada.

## II. Authors’ contributions

The authors have all significantly contributed to this work:

AJ: Conceptualization, Methodology, Data curation, Investigation, Validation, Formal analysis, visualization, Project administration, study design, Writing - original draft, Writing - review & editing, prime author.

EM: Conceptualization, Methodology, Investigation, Validation, Study design, methodology, Writing - review & editing, Project administration, Resources, Funding acquisition, Supervision.

SK: Conceptualization, Methodology, Software, Investigation, Formal analysis, Writing – review & editing

WZ: Data curation, Validation, visualization, Writing - review & editing

SG: Methodology, Software, Writing - review & editing

RA: Data curation, Validation, visualization, Writing - review & editing

AC: Data curation, Validation, visualization, Writing - review & editing

AR: Data curation, Validation, visualization, Writing - review & editing

RL: Data curation, Validation, visualization, Writing - review & editing

YL: Data curation, Validation, visualization, Writing - review & editing

YF: Data curation, Validation, visualization, Writing - review & editing

NH: Data curation, Validation, visualization, Writing - review & editing

SA: Data curation, Validation, visualization, Writing - review & editing

ZS: Data curation, Validation, visualization, Writing - review & editing

EL: Conceptualization, Methodology, Data curation, Investigation, Validation, Visualization, Writing – review & editing

RK: Conceptualization, Methodology, Investigation, Validation, Study design, methodology, Writing - review & editing, Study design, ethics approvals, senior author, corresponding author, Project administration, Resources, Funding acquisition, Supervision.

## III. Acknowledgment

N/A

## Notes

### Author Declarations

Ethics committee/IRB of The Ottawa Hospital gave ethical approval for this work.

